# Assessment of frailty in older adults: Psychometric properties of the Multidimensional Frailty Scale (MFS)

**DOI:** 10.64898/2026.01.09.26343718

**Authors:** Arantxa Gorostiaga, Joanes Lameirinhas, Igone Etxeberria, Jone Aliri

## Abstract

Frailty is increasingly recognised as a multidimensional construct. However, existing instruments assessing multidimensional frailty either show limited validity evidence or fail to adequately cover all the proposed theoretical dimensions. To address this gap, we recently developed the preliminary items of the Multidimensional Frailty Scale (MFS), a new instrument designed to assess physical, cognitive, affective, social and environmental frailty in older adults. This study sought to determine the definitive item set and to evaluate the psychometric properties of the final version. We conducted a cross-sectional observational study with 283 individuals. Participants completed the preliminary 51-item version of the MFS, together with other measures of related constructs and a sociodemographic questionnaire. Item analysis yielded a final 29-item version of the MFS. Confirmatory factor analysis supported a five-factor second-order model with excellent fit to the data. Internal consistency was high for all dimensions and the total scale. Convergent validity was supported by moderate-to-strong correlations between corresponding MFS and TFI dimensions. Similarly, the expected associations of the MFS with age, loneliness, life satisfaction and independence in activities of daily living provided validity evidence based on relations to other variables. Overall, the MFS has robust psychometric properties and constitutes a brief yet comprehensive instrument for assessing multidimensional frailty. Its hierarchical structure allows both global and domain-specific assessment, supporting more precise identification of frailty profiles. These features make the MFS suitable for use in both applied settings and research, which may contribute to earlier detection and better targeting of preventive strategies for older adults.

## 1. INTRODUCTION

Frailty is a major challenge for healthcare and social service systems worldwide, particularly in the context of ageing populations. It is generally defined as a state of vulnerability to external and internal stressors, which increases the risk of adverse health outcomes, including falls, functional limitations, cognitive disorders, institutionalisation and death (Chu et al. 2021; Fried et al. 2001; Vermeiren et al. 2016; World Health Organization 2015). Given the high prevalence of frailty and its prognostic value, the early identification of frail individuals constitutes a key element in geriatrics and gerontology, as it may help anticipate adverse health outcomes and guide the implementation of tailored interventions.

Consistent with its traditional conceptualisation as an exclusively physical condition, for many years frailty assessment focused primarily on physical indicators such as walking speed and grip strength. However, over the last decade, an increasing number of academics and practitioners have advocated for a multidimensional approach that incorporates not only physical but—at the very least— psychological and social factors also (Avgerinou et al. 2021; Cohen et al. 2023; Veronese and Pilotto 2024), as this perspective provides a broader understanding of older adults’ vulnerabilities and needs (Dury et al. 2018; Wleklik et al. 2020). Various instruments have been developed to assess multidimensional frailty, with the Tilburg Frailty Indicator (Gobbens et al. 2010) and the Comprehensive Frailty Assessment Instrument (De Witte et al. 2013) being the most widely used. Nevertheless, these instruments have shown limited validity evidence (Huang and Lam 2021; Sutton et al. 2016; Zamora-Sánchez et al. 2022) or have failed to include all the frailty dimensions proposed in the literature, such as cognitive or environmental frailty. Furthermore, the psychological frailty dimension has been inconsistently operationalised, with many instruments including both cognitive and affective indicators in it, which may compromise accuracy. It has therefore been suggested that psychological frailty should be divided into two distinct dimensions—cognitive frailty and affective frailty—to enable a more comprehensive and accurate assessment of the construct (Lameirinhas et al. 2024; Shimokihara et al. 2026).

Given the above, we deemed it necessary to develop and validate a new instrument to assess frailty from a multidimensional perspective, explicitly distinguishing between the cognitive and affective domains, while also ensuring brevity and ease of use to make it suitable for application across a range of applied settings (e.g., primary care, geriatric services and community programmes), as well as for research purposes. Accordingly, we recently began work on developing the Multidimensional Frailty Scale (MFS), a self-report measure aimed at assessing five dimensions of frailty in older adults: physical, cognitive, affective, social and environmental. Specifically, we conducted three sequential preliminary studies, described in detail elsewhere (Lameirinhas et al. 2025). In the first study, we created a draft version of the test with 83 initial items that were evaluated by a panel of 13 experts to determine their substantive and psychometric adequacy. In the second study, we tested the comprehensibility of the items through cognitive interviews with 23 older adults. Finally, we conducted a pilot study with 50 participants to preliminarily analyse item functioning and internal consistency, finding satisfactory results for both. These studies led to a refined 51-item version of the MFS, which requires further experimental validation.

Building on the version derived from the pilot study, this paper reports the results of the MFS validation study. The aims were to determine the final set of items and to evaluate the psychometric properties of the definitive version, thereby providing evidence for its reliability and validity as a multidimensional assessment instrument for frailty in older adults.

## 2. METHODS

### 2.1. Study design and ethical considerations

We conducted a cross-sectional observational study, following the most widely accepted and up-to-date standard guidelines for test development and validation (American Educational Research Association et al. 2014; Boateng et al. 2018; DeVellis and Thorpe 2021; Irwing et al. 2018).

The study complied with the principles of the Declaration of Helsinki (World Medical Association 2024) and was approved by the University of the Basque Country’s Research and Teaching Ethics Committee (reference: M10/2020/151). Written informed consent was obtained from all participants prior to data collection.

### 2.2. Participants

Based on methodological recommendations for test development and validation (e.g., Boateng et al. 2018; Ferrando and Anguiano-Carrasco 2010; Ferrando et al. 2022; Muñiz and Fonseca-Pedrero 2019), a minimum sample size of 200 participants was established prior to recruitment. Participants were recruited using a nonprobability sampling procedure. Individuals with cognitive impairment were excluded because such impairment may have compromised their understanding of the items and, consequently, the validity of their responses.

The final sample comprised 283 participants from Spain, aged 65–98 years (*M* = 76.1; *Mdn* = 75; *SD* = 9.2), of whom 174 (61.5 %) were women. Most participants were married or partnered (51.6 %), had incomplete or only primary education (59.7 %) and lived in the community (84.1 %). Detailed sociodemographic characteristics are presented in Table 1.

**Table 1.**
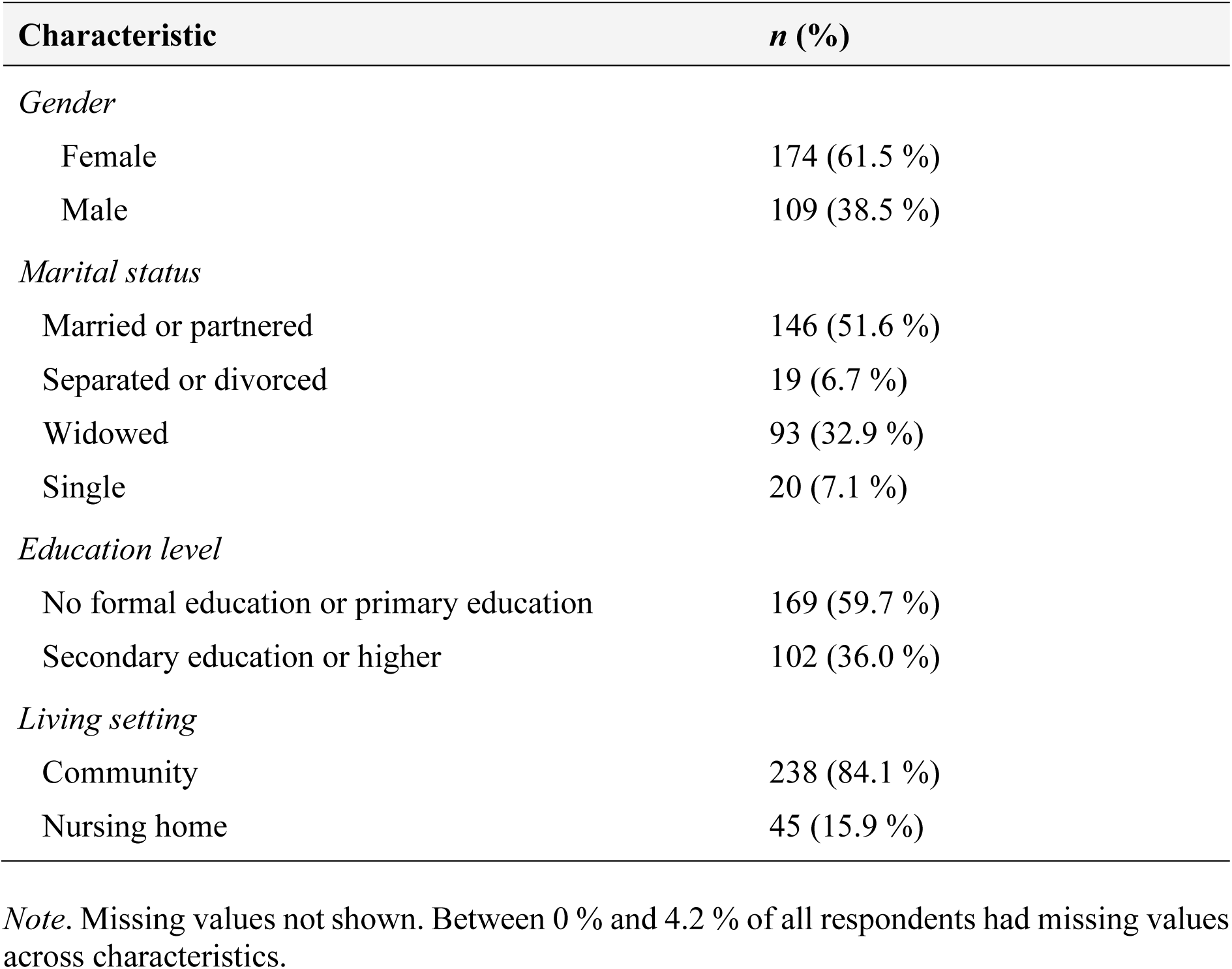
Participants’ sociodemographic characteristics.

### 2.3. Measures

The *Multidimensional Frailty Scale* (MFS). We employed the preliminary version of this self-report instrument, developed in a previous study (Lameirinhas et al. 2025), which included 51 items rated on a 4-point Likert-type scale (1 = ‘Never or almost never’; 2 = ‘Sometimes’; 3 = ‘Often’; 4 = ‘Always or almost always’) assessing physical, cognitive, affective, social and environmental frailty.

The *Tilburg Frailty Indicator* (TFI; Gobbens et al. 2010). We used the Spanish adaptation by Vrotsou et al. (2018). The TFI comprises 15 items covering physical (8 items), psychological (4 items) and social (3 items) frailty. Items are answered using either dichotomous (‘Yes’/‘No’) or 3-point (‘Yes’/‘Sometimes’/‘No’) response formats; however, all responses are later dichotomised for scoring. Total scores range from 0 to 15, with higher scores indicating greater frailty. The Cronbach’s alphas in our sample were .77 (physical), .56 (psychological), .24 (social) and .77 (overall frailty).

The *De Jong Gierveld Loneliness Scale* (DJGLS; De Jong Gierveld and Kamphuis 1985). We used the Spanish adaptation by Buz et al. (2014), which has been validated for use with older adults. The DJGLS comprises 11 items rated on a 5-point Likert-type scale (1 = ‘No!’; 2 = ‘No’; 3 = ‘More or less’; 4 = ‘Yes’; 5 = ‘Yes!’). Responses are subsequently dichotomised, with scores for each item being calculated in accordance with whether they are positively or negatively worded. Total scores range from 0 to 11, with higher scores indicating greater perceived loneliness. In the present sample, the Cronbach’s alpha was .90.

The *Satisfaction with Life Scale* (SWLS; Diener et al. 1985). We used the Spanish adaptation by Atienza et al. (2000), which was later validated for use with older adults by Pons et al. (2002). The SWLS comprises 5 items rated on a 7-point Likert-type scale (1 = ‘Totally agree’; 7 = ‘Totally disagree’). Total scores range from 7 to 35, with higher scores indicating greater life satisfaction. In the present sample, the Cronbach’s alpha was .86.

The *Barthel Index* (BI; Mahoney and Barthel 1965). We used the Spanish translation by Baztán et al. (1993), which was later validated for use with older adults by González et al. (2018). The BI assesses individuals’ level of independence in performing 10 basic activities of daily living (ADLs): feeding, bathing or showering, dressing, grooming, bowel control, bladder control, toilet use, transfers, mobility and stair climbing. Each activity is assigned a score ranging from 0 to 15. Total scores range from 0 to 100, with higher scores indicating greater independence in ADLs. In the present sample, the Cronbach’s alpha was .93.

The *Lawton and Brody Scale* (LBS; Lawton and Brody 1969). We used the Spanish adaptation by Vergara et al. (2012), which has been validated for use with older adults. The LBS assesses individuals’ level of independence in performing 8 instrumental activities of daily living (IADLs): using the telephone, shopping, preparing food, housekeeping, doing laundry, using transport, managing medication and handling finances. Although the response options vary across activities, each option is assigned a score of either 0 or 1, indicating dependence or independence, respectively. Total scores range from 0 to 8, with higher scores indicating greater independence in IADLs. In the present sample, the Cronbach’s alpha was .93.

*Sociodemographic questionnaire*. We designed an ad hoc questionnaire for this study to collect information on participants’ age, gender, marital status, education level and living setting.

### 2.4. Procedure

We contacted residential care facilities, community centres and older adult associations to recruit participants. We informed eligible individuals about the study and invited them to participate, and those who agreed provided written informed consent. Completing the questionnaires took approximately 40 minutes. For participants who were illiterate, we read the items aloud and recorded their responses in the questionnaires.

### 2.5. Data analysis

To select the final items of the MFS from the preliminary 51-item version, we calculated, for each one, the Cronbach’s alpha value if the item were deleted, the corrected item–dimension correlation and the standardised factor loading obtained from a confirmatory factor analysis with five first-order factors and one second-order factor. For the latter two indicators, items with values below .300 were considered candidates for removal (Boateng et al. 2018).

After selecting the final items, we examined the psychometric properties of the final version of the MFS using several different procedures. First, to analyse its dimensionality, we tested two confirmatory factor analysis models (a unidimensional model and a five-factor second-order model), using the weighted least squares mean and variance adjusted (WLSMV) estimator. Model fit was evaluated using the comparative fit index (CFI), the Tucker–Lewis index (TLI) and the root mean square error of approximation (RMSEA). Values above .95 for CFI and TLI were deemed to indicate good fit, as were RMSEA values below .06 (Brown 2015; Hu and Bentler 1999). Second, we assessed the internal consistency of the factors using Cronbach’s alpha, with values above .70 being considered adequate (DeVellis and Thorpe 2021). Third, to obtain evidence of convergent validity, we calculated Spearman’s correlations between corresponding dimensions of the MFS and the TFI. Following Cohen’s criteria (1988), the cutoff points for effect sizes were .10 (small), .30 (moderate) and .50 (large). Finally, to examine validity evidence based on relations to other variables, we calculated Spearman’s correlations of the total score and selected dimensions of the MFS with age, loneliness, life satisfaction and independence in ADLs and IADLs. Drawing from previous research, we hypothesised that age would correlate positively with physical frailty, loneliness would correlate positively with affective and social frailty, life satisfaction would correlate negatively with affective and social frailty, and independence in ADLs and IADLs would correlate negatively with physical frailty. We performed the analyses using Mplus (version 8.1) and IBM SPSS Statistics (version 29.0).

## 3. RESULTS

### 3.1. Item selection

Supplementary Table 1 shows, for each item, the Cronbach’s alpha value if item deleted, the corrected item–dimension correlation and the standardised factor loading. Based on these indices, the six best-performing items were retained for each dimension, except for the environmental dimension, for which only five items were selected, resulting in a total of 29 items. Content validity was also taken into account throughout the decision-making process, drawing on evidence from the literature and the expert consultation conducted during the initial stages of the instrument’s validation, to ensure that the retained items adequately represented the conceptual domain of each dimension.

The final version of the MFS, along with an English translation of the items, is provided in Supplementary Table 2.

### 3.2. Dimensionality and internal consistency

Table 2 presents the fit indices of the two tested models. Only the five-factor second-order model—whose structure is displayed in Figure 1—was found to have good fit. As reported in Supplementary Table 3, the Cronbach’s alpha coefficients for the five dimensions ranged from .836 to .937, and the Cronbach’s alpha for the total scale was .931. The corrected item–dimension correlations ranged from .482 to .903, and the standardised factor loadings of the items on their respective dimensions ranged from .779 to .972 (except for item AF-1, which had a loading of .570).

**Figure 1.**
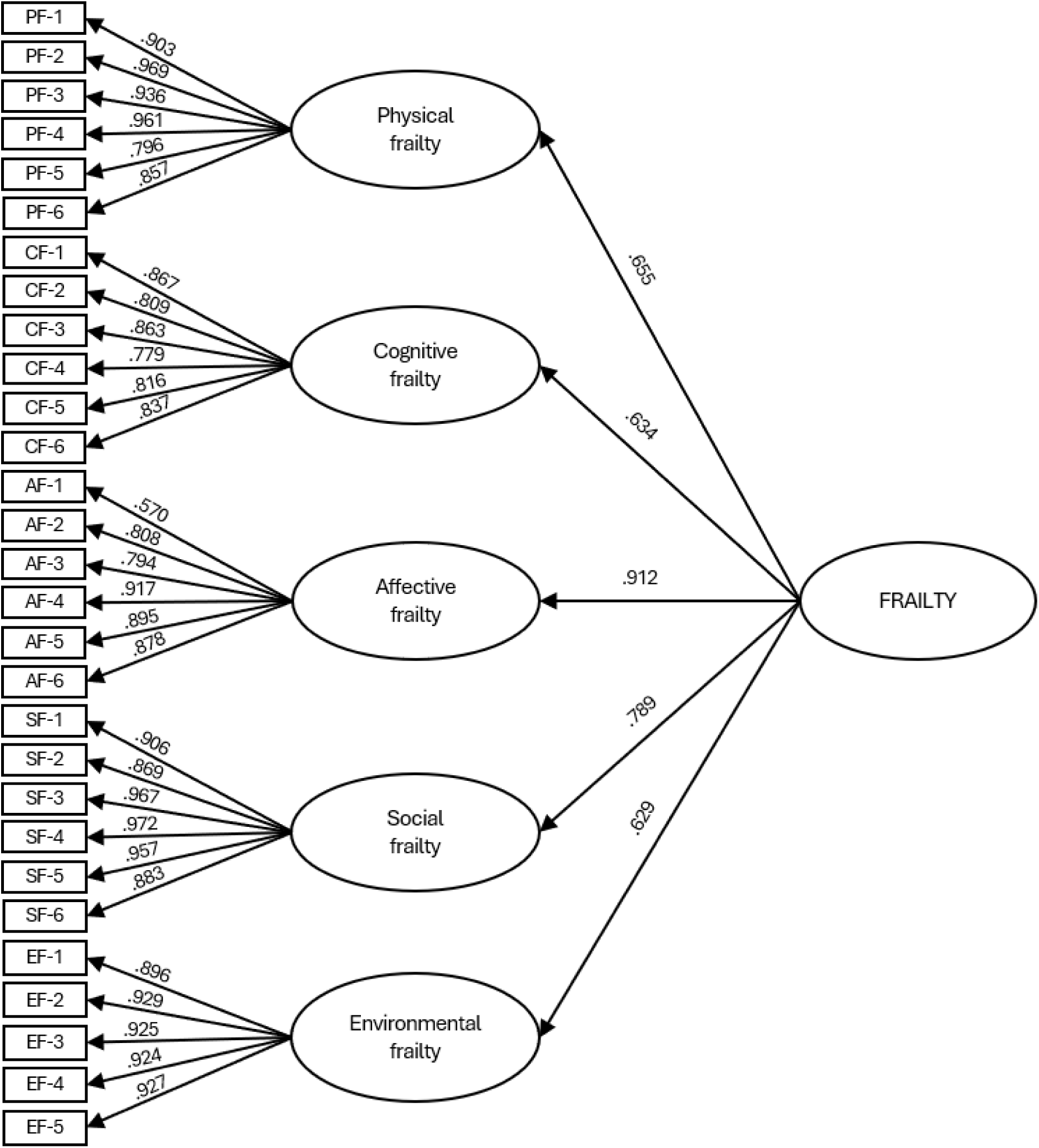
Confirmatory factor analysis of the five-factor second-order model of the Multidimensional Frailty Scale (MFS)

**Table 2.**
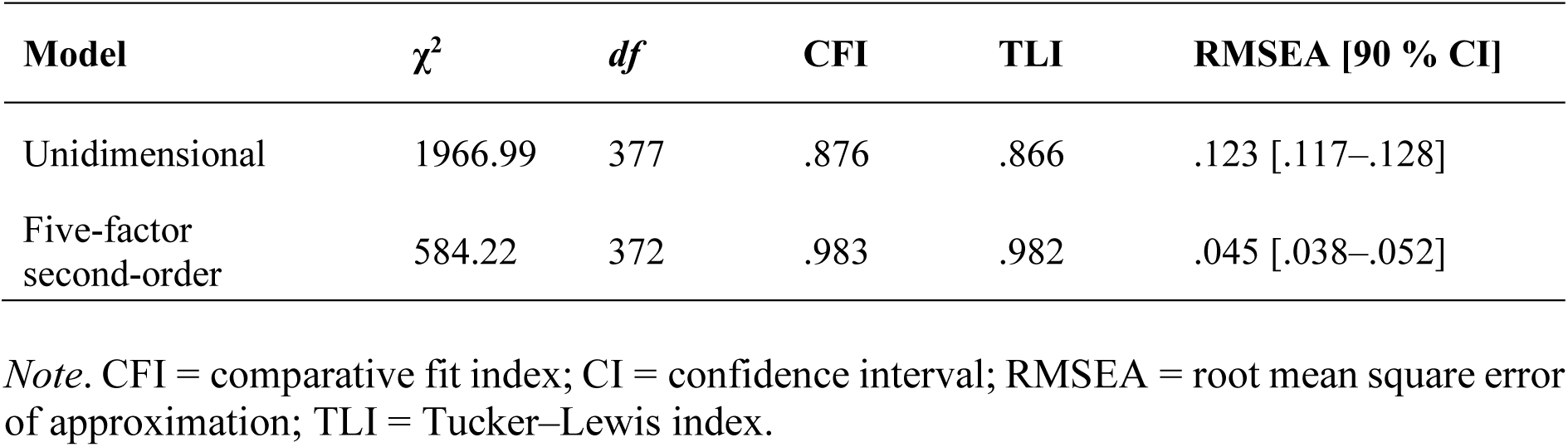
Fit indices of the tested confirmatory factor analysis models.

| Model | $\chi^2$ | *df* | CFI | TLI | RMSEA [90 % CI] |
| --- | --- | --- | --- | --- | --- |
| Unidimensional | 1966.99 | 377 | .876 | .866 | .123 [.117–.128] |
| Five-factor second-order | 584.22 | 372 | .983 | .982 | .045 [.038–.052] |

*Note.* CFI = comparative fit index; CI = confidence interval; RMSEA = root mean square error of approximation; TLI = Tucker–Lewis index.

### 3.3. Convergent validity

Table 3 shows the Spearman correlations between the MFS and the TFI. As expected, the strongest correlations were observed between subscales assessing similar dimensions (e.g., physical frailty, *r* = .809). Likewise, the correlation between the psychological dimension of the TFI and the affective dimension of the MFS was stronger than the one between the psychological dimension of the TFI and the cognitive dimension of the MFS.

**Table 3.**
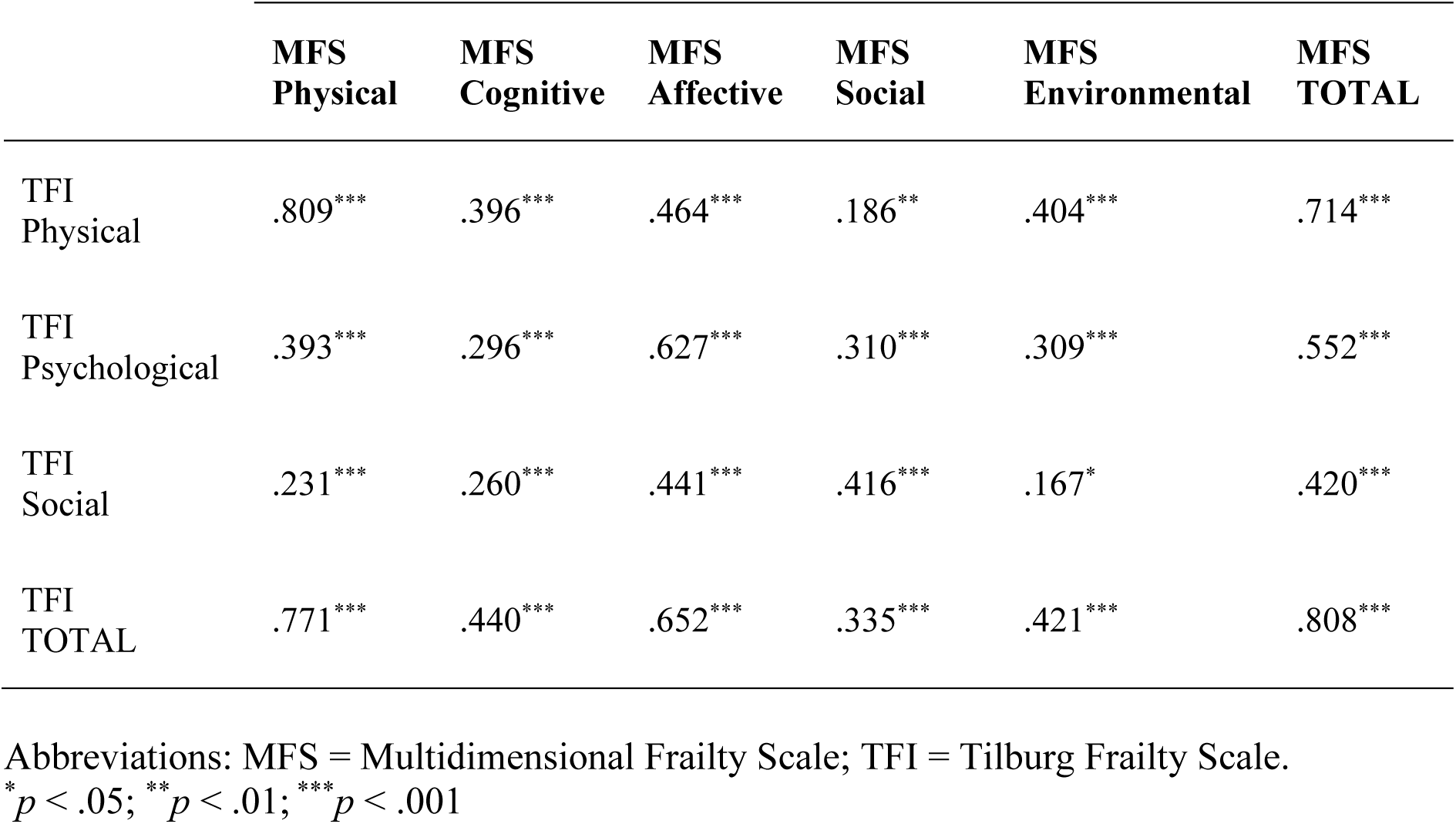
Spearman correlations between the dimensions of the Multidimensional Frailty Scale (MFS) and the Tilburg Frailty Indicator (TFI)

|  | **MFS**<br>**Physical** | **MFS**<br>**Cognitive** | **MFS**<br>**Affective** | **MFS**<br>**Social** | **MFS**<br>**Environmental** | **MFS**<br>**TOTAL** |
| --- | --- | --- | --- | --- | --- | --- |
| TFI<br>Physical | .809*** | .396*** | .464*** | .186** | .404*** | .714*** |
| TFI<br>Psychological | .393*** | .296*** | .627*** | .310*** | .309*** | .552*** |
| TFI<br>Social | .231*** | .260*** | .441*** | .416*** | .167* | .420*** |
| TFI<br>TOTAL | .771*** | .440*** | .652*** | .335*** | .421*** | .808*** |

Abbreviations: MFS = Multidimensional Frailty Scale; TFI = Tilburg Frailty Scale.
\* $p < .05$ ; \*\* $p < .01$ ; \*\*\* $p < .001$

### 3.4. Validity evidence based on relations to other variables

Table 4 presents the Spearman correlations of the total score and selected dimensions of the MFS with age, loneliness, life satisfaction and independence in ADLs and IADLs. Age was found to correlate moderately and positively with physical frailty and weakly and positively with affective frailty, whereas its correlation with social frailty was negligible and non-significant. Loneliness was found to correlate weakly and positively with physical frailty, and strongly and positively with affective and social frailty. Life satisfaction correlated weakly and negatively with physical and social frailty and moderately and negatively with affective frailty. Independence in ADLs and IADLs correlated strongly and negatively with physical frailty and moderately and negatively with affective frailty, whereas their correlations with social frailty were weak and non-significant. Regarding the total MFS score, moderate positive correlations were observed with age and loneliness, a moderate negative correlation was observed with life satisfaction, and strong negative correlations were observed with independence in both ADLs and IADLs.

**Table 4.** Spearman correlations of the total score and selected dimensions of the Multidimensional Frailty Scale (MFS) with age, loneliness, life satisfaction and independence in basic and instrumental activities of daily living.

|  | **MFS**<br>**Physical** | **MFS**<br>**Affective** | **MFS**<br>**Social** | **MFS**<br>**TOTAL** |
| --- | --- | --- | --- | --- |
| Age | .435*** | .172** | .098 | .388*** |
| Loneliness | .231*** | .543*** | .573*** | .473*** |
| Life satisfaction | -.183** | -.421*** | -.284*** | -.313*** |
| Independence in ADLs | -.732*** | -.335*** | -.112 | -.629*** |
| Independence in IADLs | -.732*** | -.328*** | -.126 | -.639*** |

*Note.* ADLs = Basic activities of daily living; IADLs = Instrumental activities of daily living; MFS = Multidimensional Frailty Scale.
\*\* $p < .01$ ; \*\*\* $p < .001$

## 4. DISCUSSION

The present study aimed to determine the final set of items of the MFS and to evaluate its psychometric properties in individuals aged 65 and over. The item analysis allowed us to identify the best-performing items while keeping the scale at a reasonable length, with the goal of creating an instrument suitable for use across a range of applied settings (e.g., primary care, geriatric services and community programmes), as well as for research purposes. The final version of the MFS comprises 29 items and allows frailty to be assessed both globally and across its physical, cognitive, affective, social and environmental dimensions in approximately five minutes.

Regarding its dimensionality, the confirmatory factor analysis indicated that the MFS consists of five first-order dimensions (physical, cognitive, affective, social and environmental frailty) and a second-order factor representing overall frailty. This hierarchical structure enables both the assessment of each specific dimension and the assessment of overall frailty. This is a notable strength of the instrument, as few existing multidimensional frailty measures examine or support a second-order factor structure of this kind that enables a dual-level assessment.

The internal consistency indices were adequate and the correlations with the TFI provided evidence of good convergent validity. As expected, moderate-to-strong correlations emerged between analogous dimensions of the two instruments. Also as expected, the correlation between the cognitive dimension of the MFS and the psychological dimension of the TFI was weak, which is consistent with the fact that the latter predominantly captures affective components. These findings highlight the importance of assessing cognitive and affective indicators of frailty separately, as suggested in previous studies (Facal et al. 2019; Lameirinhas et al. 2024; Shimokihara et al. 2026).

In terms of validity evidence based on relations to other variables, as hypothesised and consistently with the extant literature (e.g., Chen et al. 2021; Collard et al. 2012; Liu et al. 2020; Merchant et al. 2017), a moderate positive correlation was found between age and physical frailty. This relationship may be explained by age-related loss of muscle mass, which can reduce strength and physical endurance, thereby increasing the risk of physical frailty and, subsequently, disability (Fielding 2015; Phillips et al. 2017; Zhou et al. 2024). In contrast, and again consistently with that reported by previous studies (e.g., Kasa et al. 2024; van Assen et al. 2016; Ye et al. 2021), only a weak positive correlation was observed between age and affective frailty, along with a negligible, non-significant correlation with social frailty. These findings suggest that, given the well-established relationship between age and physical functioning, age may be more closely related to the physical dimension of frailty than to its affective and social dimensions, in which other factors, such as social support and major life events (e.g., widowhood), may play a greater role. The findings for the remaining variables were also consistent with previous research: loneliness was positively associated with both affective and social frailty, with large effect sizes (e.g., Altintas et al. 2023; Klesiora et al. 2024; Li et al. 2024; Ye et al. 2024); life satisfaction was negatively associated with these dimensions, with a moderate effect size for affective frailty and a small effect size for social frailty (e.g., Duppen et al. 2019; Hoeyberghs et al. 2018; Jeong and Chang 2023; Ko and Jung 2021); and independence in ADLs and IADLs was strongly and negatively associated with physical frailty (e.g., Chen et al. 2021; Coelho et al. 2015; Doñate-Martínez et al. 2022; Gobbens et al. 2014; Kojima 2017; Urrunaga-Pastor et al. 2024).

Overall, the findings of the present study provide evidence of the reliability and validity of the MFS as a multidimensional assessment instrument for frailty in older adults—something for which professionals are calling with increasing frequency (Avgerinou et al. 2021; Cohen et al. 2023; Veronese and Pilotto 2024). It is also important to note that this paper builds on a sequence of preliminary research studies (Lameirinhas et al. 2025) that provide additional validity evidence of the MFS: a comprehensive literature review and an expert consultation (validity evidence based on test content), a cognitive pretest (validity evidence based on response processes) and a pilot study (preliminary validity evidence based on internal structure).

Beyond its psychometric properties, the MFS offers several practical advantages. First, it encompasses all the dimensions of frailty proposed in the literature—physical, cognitive, affective, social and environmental—whereas existing multidimensional instruments generally cover only some of these domains. Importantly, the MFS assesses cognitive and affective indicators of frailty within separate dimensions, rather than including them indiscriminately within a broader dimension labelled ‘psychological frailty’, as is common in existing multidimensional frailty instruments. This distinction provides a more accurate and nuanced representation of frailty by recognising cognitive and affective vulnerability as distinct dimensions that may follow different trajectories, be influenced by different risk factors and affect individuals in different ways (Facal et al. 2019; Lameirinhas et al. 2024). Moreover, unlike instruments that yield only an overall frailty score (e.g., frailty indices), the MFS provides both a global frailty score and domain-specific scores. The global score provides a concise, integrated measure of an individual’s overall frailty. In turn, the domain-specific scores provide a more detailed characterisation of individual frailty profiles by identifying areas of particular vulnerability, and may therefore help inform the planning of tailored preventive or intervention strategies. Another practical advantage is the standardised nature of the MFS. Many frailty assessment instruments, particularly frailty indices, are developed ad hoc, with the variables included varying from one study to another. In contrast, the MFS comprises a fixed set of items and uses a standardised scoring procedure, which may facilitate comparisons across studies and populations and contribute to the accumulation of more consistent and comparable evidence on multidimensional frailty. Finally, owing to its brevity and ease of administration, the MFS may be readily used in practical health and social care settings, as well as in research.

However, some limitations must be acknowledged. First, the MFS relies on self-reported data, which may introduce response biases related to literacy, introspective ability, self-awareness and social desirability. Second, the use of a nonprobability sampling method may reduce the representativeness of the sample and, consequently, restrict the generalisability of the findings. Finally, data could not be collected to assess test–retest reliability, an aspect that should be examined in future studies.

Despite these limitations, however, we believe that the MFS, with its multidimensional perspective on frailty, holds great potential for both researchers (by providing a psychometrically sound tool for the study of multidimensional frailty) and practitioners (by supporting more accurate identification of frailty). Such a comprehensive assessment may facilitate early detection and better targeting of preventive and care strategies, thereby helping to prevent adverse health outcomes and safeguard the well-being of older adults. This is of paramount importance given the steady growth of the older population and the risk of frailty becoming a prevalent condition within this demographic.

## Data Availability

All data produced in the present study are available upon reasonable request to the authors.

