## Supplemental material for "Assessment of frailty in older adults: Psychometric properties of the Multidimensional Frailty Scale (MFS)"

This document contains the supplementary materials of the article entitled "Assessment of frailty in older adults: Psychometric properties of the Multidimensional Frailty Scale (MFS)", authored by Arantxa Gorostiaga, Joanes Lameirinhas, Igone Etxeberria, and Jone Aliri. The index of the supplementary materials is provided below.


| SUPPLEMENTARY TABLE 1 | |
|---|---|
| Participants' sociodemographic characteristics | 2 |

| SUPPLEMENTARY TABLE 2 | |
|---|---|
| Cronbach's alpha if item deleted, corrected item–dimension correlations, and standardized factor loadings for the preliminary 51-item version of the Multidimensional Frailty Scale (MFS) | 3 |

| SUPPLEMENTARY TABLE 3 | |
|---|---|
| Spanish items of the MFS and their translation into English | 5 |

| SUPPLEMENTARY TABLE 4 | |
|---|---|
| Cronbach's alpha if item deleted, corrected item–dimension correlations, and standardized factor loadings for the final 29-item version of the Multidimensional Frailty Scale (MFS) | 8 |

**Supplementary Table S1.** Participants' sociodemographic characteristics

| **Characteristic** | ***n* (%)** |
|---|---|
| Gender | |
| Female | 174 (61.5%) |
| Male | 109 (38.5%) |
| Marital status | |
| Married or partnered | 146 (51.6%) |
| Separated or divorced | 19 (6.7%) |
| Widowed | 93 (32.9%) |
| Single | 20 (7.1%) |
| Education level | |
| No formal education or primary education | 169 (59.7%) |
| Secondary education or higher | 102 (36.0%) |
| Living setting | |
| Community | 238 (84.1%) |
| Nursing home | 45 (15.9%) |

*Note*: Missing values not shown. Between 0% and 4.2% of all respondents had missing values across characteristics.

**Supplementary Table S2.** Cronbach's alpha if item deleted, corrected item–dimension correlations, and standardized factor loadings for the preliminary 51-item version of the Multidimensional Frailty Scale (MFS)

| Item number in the 51-item version | Cronbach's alpha if item deleted | Corrected item–dimension correlation | Standardized factor loading |
|---|---|---|---|
| PHYSICAL FRAILTY (Cronbach's alpha = .930) | | | |
| PF_1* | .927 | .664 | .812 |
| PF_2 | .918 | .812 | .902 |
| PF_3 | .913 | .878 | .964 |
| PF_4 | .917 | .831 | .924 |
| PF_5 | .913 | .886 | .961 |
| PF_6 | .927 | .662 | .782 |
| PF_7* | .927 | .669 | .744 |
| PF_8 | .919 | .790 | .873 |
| PF_9* | .936 | .493 | .603 |
| COGNITIVE FRAILTY (Cronbach's alpha = .909) | | | |
| CF_1 | .900 | .661 | .844 |
| CF_2* | .898 | .689 | .802 |
| CF_3 | .897 | .707 | .815 |
| CF_4 | .898 | .695 | .849 |
| CF_5 | .900 | .665 | .828 |
| CF_6* | .904 | .574 | .713 |
| CF_7* | .900 | .655 | .771 |
| CF_8 | .896 | .725 | .855 |
| CF_9* | .902 | .626 | .704 |
| CF_10* | .908 | .503 | .570 |
| CF_11 | .898 | .689 | .834 |
| AFFECTIVE FRAILTY (Cronbach's alpha = .933) | | | |
| AF_1 | .936 | .507 | .608 |
| AF_2* | .932 | .571 | .721 |
| AF_3* | .924 | .777 | .849 |
| AF_4 | .927 | .712 | .837 |
| AF_5 | .929 | .660 | .770 |
| AF_6 | .923 | .788 | .918 |
| AF_7 | .926 | .736 | .882 |
| AF_8* | .923 | .787 | .886 |
| AF_9 | .923 | .800 | .943 |
| AF_10* | .923 | .807 | .943 |
| AF_11* | .924 | .770 | .901 |

| | | | |
|---|---|---|---|
| **SOCIAL FRAILTY (Cronbach's alpha = .889)** | | | |
| SF_1 | .876 | .677 | .887 |
| SF_2 | .875 | .671 | .833 |
| SF_3 | .870 | .782 | .964 |
| SF_4 | .870 | .777 | .967 |
| SF_5 | .868 | .792 | .945 |
| SF_6* | .883 | .554 | .750 |
| SF_7* | .893 | .440 | .676 |
| SF_8* | .902 | .449 | .750 |
| SF_9 | .872 | .740 | .891 |
| SF_10* | .881 | .577 | .764 |
| SF_11* | .877 | .658 | .804 |
| **ENVIRONMENTAL FRAILTY (Cronbach's alpha = .778)** | | | |
| EF_1 | .753 | .545 | .884 |
| EF_2 | .740 | .573 | .922 |
| EF_3 | .750 | .545 | .906 |
| EF_4 | .734 | .602 | .882 |
| EF_5 | .728 | .636 | .915 |
| EF_6* | .766 | .409 | .563 |
| EF_7* | .782 | .353 | .548 |
| EF_8* | .782 | .257 | .869 |
| EF_9* | .770 | .360 | .410 |

*Note*. Eliminated items are marked with an asterisk (*).

**Supplementary Table S3.** English-translated MFS items

| | |
|---|---|
| **PHYSICAL FRAILTY** | |
| PF_1 | I find it difficult to carry out daily activities because I get tired easily (for example, cleaning the house, grocery shopping, going up or down stairs, taking a walk…). |
| PF_2 | My physical condition makes it difficult for me to carry out daily activities (for example, cleaning the house, carrying grocery bags, going up or down stairs, taking a walk…). |
| PF_3 | I find it difficult to carry out daily activities due to problems maintaining my balance (for example, cleaning the house, carrying grocery bags, going up or down stairs, taking a walk…). |
| PF_4 | I find it difficult to carry out daily activities because I have trouble walking (for example, cleaning the house, grocery shopping, going up or down stairs, taking a walk…). |
| PF_5 | I find it difficult to carry out daily activities because I have little strength in my hands and arms (for example, cleaning the house, opening containers, carrying grocery bags…). |
| PF_6 | I need to support myself on something (for example, a cane, a table, or an armrest) to stand up from a chair. |
| **COGNITIVE FRAILTY** | |
| CF_1 | When talking to others, I have trouble finding the words I want to say. |
| CF_2 | Words feel “on the tip of my tongue” (for example, when referring to an object or a person) |
| CF_3 | When talking to others, I find it difficult to express clearly what I want to say. |
| CF_4 | I have difficulty concentrating (for example, during conversations, reading, or watching television). |
| CF_5 | I have memory problems. |
| CF_6 | I have difficulty remembering things that happened recently (for example, a conversation from the previous day or what I ate during the day). |

| AFFECTIVE FRAILTY | |
|---|---|
| AF_1 | I feel nervous or worried. |
| AF_2 | I feel lonely (even when I am surrounded by others). |
| AF_3 | I feel that I am losing the confidence I once had in myself. |
| AF_4 | I feel that I am losing control of my life. |
| AF_5 | I feel useless. |
| AF_6 | I feel that my life is empty. |
| **SOCIAL FRAILTY** | |
| SF_1 | I lack people who are willing to help me when I need it (friends, relatives, partner, neighbors...). |
| SF_2 | I lack trusted people around me with whom I can talk about everyday problems (friends, relatives, partner, neighbors...). |
| SF_3 | I lack people who care about me (friends, relatives, partner, neighbors...). |
| SF_4 | I lack people who appreciate me (friends, relatives, partner, neighbors...). |
| SF_5 | I lack people who make me feel loved and protected (friends, relatives, partner, neighbors...). |
| SF_6 | I lack social relationships that are important to me (friends, relatives, partner, neighbors...). |

| ENVIRONMENTAL FRAILTY | |
|---|---|
| EF_1 | I find it difficult to move around my home* due to architectural barriers (for example, limited space, narrow hallways, uneven surfaces, poor lighting…). |
| EF_2 | I have difficulty entering or leaving my home* due to architectural barriers (for example, stairs and/or lack of an elevator…). |
| EF_3 | I stay at home* without going outside because of the architectural barriers in my home* (for example, stairs, lack of an elevator, living far from town**…). |
| EF_4 | I have difficulty moving around my neighborhood and/or town** due to architectural barriers (for example, steep slopes, poorly maintained sidewalks, living in an isolated or poorly connected area…). |
| EF_5 | I find it difficult to reach local shops and services in my neighborhood and/or town** due to architectural barriers (for example, steep slopes, poorly maintained sidewalks, living in an isolated or poorly connected area…). |

* If the MFS is administered to a person living in a care facility, it is recommended to use the word “facility” instead of “home”.

** If the MFS is administered to a person living in a city, it is recommended to use the word “city” instead of “town”.

**Supplementary Table S4.** Cronbach's alpha if item deleted, corrected item–dimension correlations, and standardized factor loadings for the final 29-item version of the Multidimensional Frailty Scale (MFS)

| Item number in the 51-item version | Item number in the 29-item version | Cronbach's alpha if item deleted | Corrected item–dimension correlation | Standardized factor loading |
|---|---|---|---|---|
| PHYSICAL FRAILTY (Cronbach's alpha = .937) | | | | |
| PF_2 | PF_1 | .927 | .808 | .903 |
| PF_3 | PF_2 | .914 | .903 | .969 |
| PF_4 | PF_3 | .919 | .866 | .936 |
| PF_5 | PF_4 | .917 | .880 | .961 |
| PF_6 | PF_5 | .941 | .677 | .796 |
| PF_8 | PF_6 | .934 | .760 | .857 |
| COGNITIVE FRAILTY (Cronbach's alpha = .874) | | | | |
| CF_1 | CF_1 | .846 | .715 | .867 |
| CF_3 | CF_2 | .849 | .697 | .809 |
| CF_4 | CF_3 | .847 | .711 | .863 |
| CF_5 | CF_4 | .866 | .597 | .779 |
| CF_8 | CF_5 | .855 | .664 | .816 |
| CF_11 | CF_6 | .852 | .680 | .837 |
| AFFECTIVE FRAILTY (Cronbach's alpha = .871) | | | | |
| AF_1 | AF_1 | .882 | .482 | .570 |
| AF_4 | AF_2 | .852 | .657 | .808 |
| AF_5 | AF_3 | .848 | .681 | .794 |
| AF_6 | AF_4 | .829 | .783 | .917 |
| AF_7 | AF_5 | .838 | .739 | .895 |
| AF_9 | AF_6 | .844 | .702 | .878 |
| SOCIAL FRAILTY (Cronbach's alpha = .931) | | | | |
| SF_1 | SF_1 | .923 | .759 | .906 |
| SF_2 | SF_2 | .923 | .765 | .869 |
| SF_3 | SF_3 | .912 | .849 | .967 |
| SF_4 | SF_4 | .913 | .839 | .972 |
| SF_5 | SF_5 | .911 | .854 | .957 |
| SF_9 | SF_6 | .927 | .726 | .883 |
| ENVIRONMENTAL FRAILTY (Cronbach's alpha = .836) | | | | |
| EF_1 | EF_1 | .827 | .571 | .896 |
| EF_2 | EF_2 | .785 | .701 | .929 |
| EF_3 | EF_3 | .793 | .708 | .925 |
| EF_4 | EF_4 | .830 | .567 | .924 |
| EF_5 | EF_5 | .774 | .738 | .927 |

*Note*: The total Cronbach's alpha value was .931.
